# International Completeness of Death Registration 2015-2019

**DOI:** 10.1101/2021.08.12.21261978

**Authors:** Ariel Karlinsky

## Abstract

Death registration completeness, the share of deaths captured by countries’ vital registration systems, vary substantially across countries. Estimates of completeness, even recent ones, are outdated or contradictory for many countries. This paper presents the International Completeness of Death Registration (ICDR) dataset. ICDR contains the annual amount of deaths registered in 181 countries’ vital registration systems from 2015 to 2019. These counts allow derivation of the most up-to-date and consistent estimates of death registration completeness.

## 1 Introduction

If a death occurs, how likely is it to be registered? the answer varies considerably across regions, economic development level and countries. Reliable information on deaths and completeness of death registration is vital for countries, international organizations and civil society as it informs their decisions in all venues, especially health and population matters. Without sufficient and well-understood data, the effect of policies and interventions cannot be understood properly. Moreover, for countries to understand if they are on the path to achieve many of the Sustainable Development Goals (SDG 2020) such as mortality reduction and fighting communicable diseases, reliable monitoring systems must be established and their performance tested. The importance of civil vital registration systems was further emphasized in SDG goal 17.19.2 which states that by 2030 countries should achieve “100% birth registration and 80% death registration” (WHO, 2017). Moreover, as Makinde et al. (2020) note, 14 SDG indicators are reliant on civil vital registration systems. WHO (2017) further emphasizes that the preferred source to check progress of 17.19.2 is “Civil Vital Registration Systems”, followed by Censuses. The recent SCORE assessment tools (WHO, 2021) strengthened this by including “Full Birth And Death Registration” as one of it’s main indicators.

Completeness of death registration is estimated by dividing the number of registered deaths in a given period (such as year) by the number of expected deaths. With the number of expected deaths in each country-year derived using various information sources and methods such as censuses, surveys and vital registration system (see Rao et al. (2020) for a recent review). The most prominent international sources for the number of expected annual deaths are the United Nations’ World Population Prospects (2019), Institute of Health Metrics and Evaluation’s Global Burden of Disease (2020) and the World Health Organization Global Health Estimates (2021) which utilize a wide variety of methods to estimate annual expected deaths for a wide variety of countries. Based on these and other sources, several recent studies have estimated the completeness of death registration in several countries (Mikkelsen et al., 2015; Adair and Lopez, 2018; Johnson et al., 2021).

Internationally, the two most widely used sets of completeness of death registration estimates are the UN Statistical Division’s Demographic Yearbook (2021) and IHME’s Global Burden of Disease (2020). However, some UNSD (2021) completeness’s estimates are opaque, listed at “Less than 90%” (is it 40, 50 or 75% complete)? Additionally, in some countries, these datasets’ estimates are outdated. For example, in Bolivia, the UNSD (2021) estimate is from 2000 and GBD (2020) is from 2003. In other cases, the latest estimate may be fairly recent (2015 for Peru), yet it may be outdated due to significant reforms since then such as Peru’s modernization of its’ death registration system (Vargas-Herrera et al., 2018). Some of the completeness estimates are contradictory, with large differences between sources. For example, UNSD (2021) estimate the completeness of death registration in Lebanon from 2007 at “90% or more” while GBD (2020) contains no estimate for it at all.

This study presents the International Completeness of Death Registration (ICDR) dataset. ICDR was created by hand-collecting death registration data from 181 countries between 2015 to 2019 to provide the most up-to-date and reliable estimates of the completeness of death registration in one single source. ICDR is freely and publicly available at https://github.com/akarlinsky/death_registration.

## 2 Materials and Methods

In order to construct the ICDR dataset, we used several sources. First, we build our previous work, the Mortality Dataset (Karlinsky and Kobak, 2021) which contains weekly or monthly death counts from 122 countries and territories between 2015 to 2022. The data from the World Mortality Dataset (henceforth: WMD) are official counts published or provided by the responsible institutions in all countries: national statistics offices, civil registries, ministries of health, etc. Henceforth referred to as NSOs. A detailed list of sources for each country in WMD is available at https://github.com/akarlinsky/world_mortality. Inclusion criteria in WMD require some information to be available for 2020 and at least monthly time format. As this paper focuses on the 2015-2019 period, data from WMD was supplemented by official registered number of deaths, hand-collected from 64 countries for this period. Similarly to the WMD, the sources for most additional countries are official reports from the national NSOs. Additionally, we obtained death registration data for some countries from the CRVS midterm questionnaire, UNSD yearbook, the UN’s special collection of deaths by month, WHO’s Mortality Database and the research literature (UNESCAP, 2021; UNSD, 2021; UNDATA, 2021; WHO, 2021; Zeng et al., 2020; Stoneburner and Greenwell, 2017). The extensive list of sources for all countries not from the WMD is given in the References section.

If information for some country was found in more than one source, priority was given to figures published by the country itself or with a more direct attribution. For example, In Bangladesh 2018 the number of deaths in the UNSD yearbook is over 820 thousand, but this is contradicted by the figure in the CRVS midterm questionnaire which is more plausible at about 196 thousand.^1^

The annual amount of registered deaths in each country in 2015 to 2019 were contrasted with annual expected number of deaths. As discussed in the introduction, there are 3 prominent international datasets containing estimates of total/expected deaths by country-year: World Population Prospects (WPP 2019)^2^, Global Burden of Disease (GBD 2020) and Global Health Estimates (GHE 2021). The estimates in these sources are derived from a variety of sources using many methods, a detailed discussion of which is outside the scope of this work. It is important to note that the three different estimates are mostly similar, especially when vital registration systems are complete. However, the estimates sometimes differ significantly. For example, the estimate of total deaths in the United Arab Emirates in 2019 is given as 14869 (WPP), 20932 (GHE) and 29113 (GHE). Such large disagreements reflect the fact that the underlying information in such countries might be problematic and differences are driven both by modelling assumptions and methodology and selection of data sources. In order to limit the effect of such disagreements on our estimates, our final estimate for expected number of deaths in each year was obtained as the mean of the three estimates. However, the publicly availability of the ICDR dataset allows researchers to use any one of these estimates, some other function thereof or another estimate of expected deaths entirely. See section 3 for further discussion on this.

A few small changes were conducted to the registration data in order to merge it with the number of expected deaths: WMD provides separate estimates for Transnistria and Kosovo, while the three sources of expected number deaths count them as part of Moldova and Serbia, respectively. For this study, we joined them so that the completeness rate for Moldova and Serbia would not be downward-biased.^3^ The territories of Faroe Islands (Denmark), Gibraltar (UK) and the country of Liechtenstein were not available in either of the three expected deaths datasets and were removed. After these modifications, we provide data for a total of 181 countries: 117 from WMD and 64 from the additional sources discussed above.

## 3 Results

Our main result is the death registration completeness rate for each country, defined as:

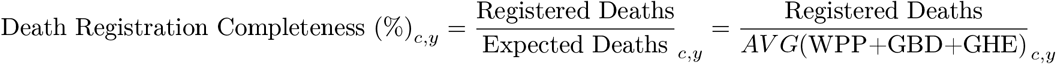

With *c* denoting country and *t* denoting year. The full list of the 181 ICDR countries and the death registration completeness estimates are provided in https://github.com/akarlinsky/death_registration along with the underlying values for the number of registered and expected deaths. It is available in a machine readable, longitudinal format at https://github.com/akarlinsky/death_registration. Figure 1 shows a map of the completeness rate in the latest available year for each country.

**Figure 1:**
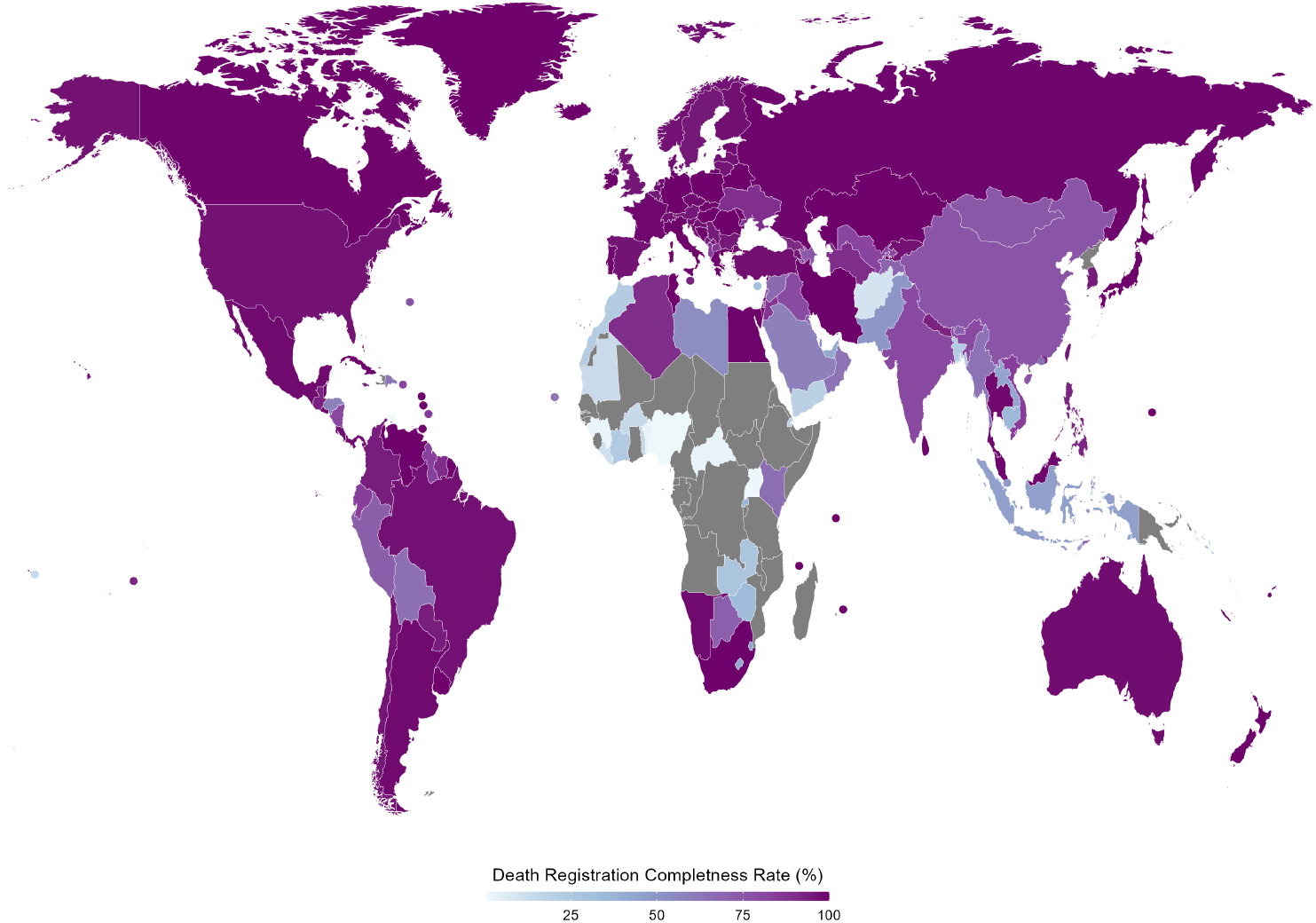
Death Registration Completeness by country. Small countries and territories are shown with circles. Grey denotes no death registration data was located.

In order to better understand our results, it’s useful to contrast them with previously reported estimates of death coverage from UNSD (2021) and GBD (2020). Table 1 presents this comparison for some selected countries.

**Table 1:**
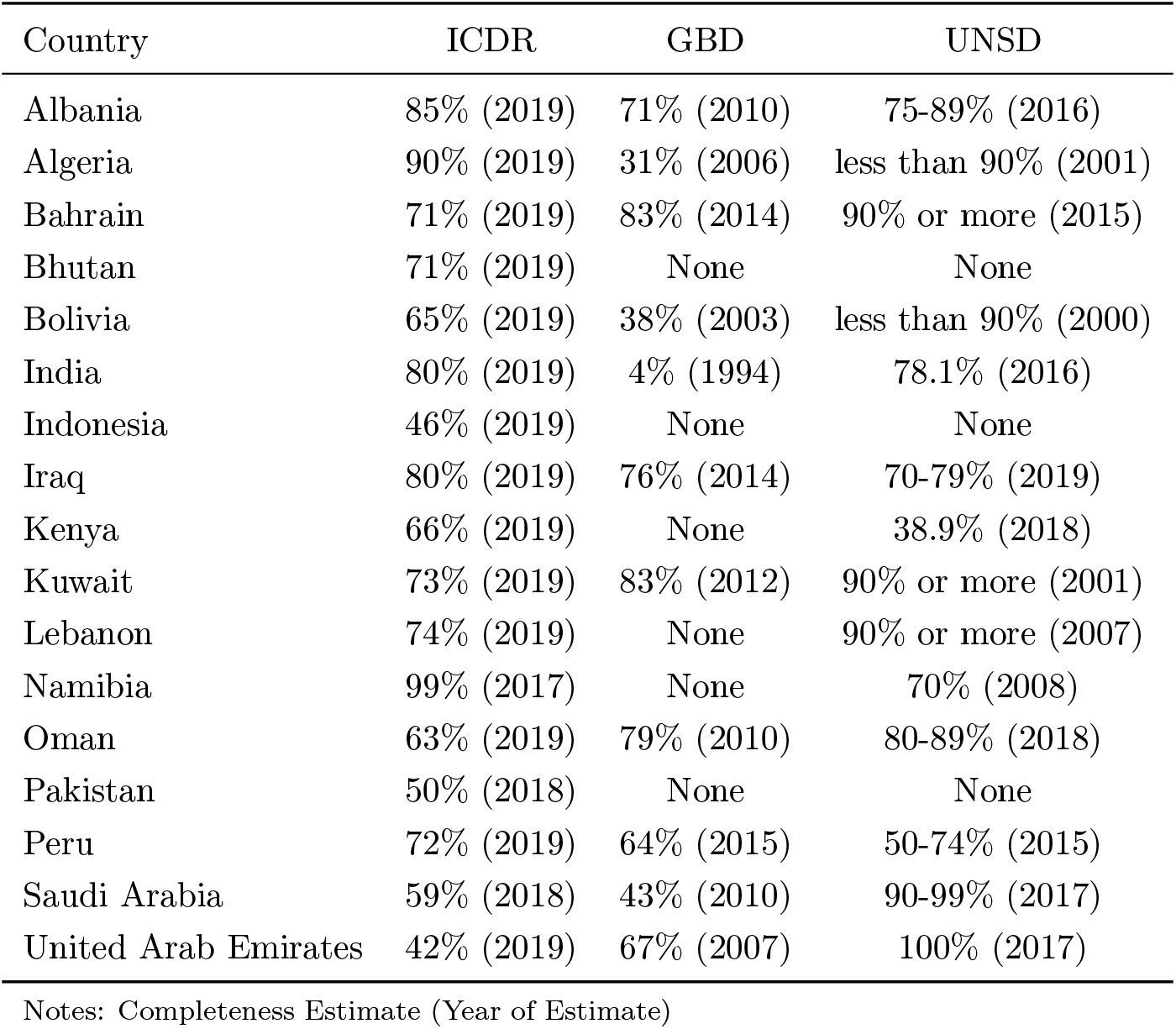
Death Registration Completeness: Selected Countries ICDR Compared to GBD and UNSD

First and foremost, ICDR covers 21 countries that neither GBD nor UNSD contain estimates for, such as Indonesia, Ivory Coast and Bhutan. Second, ICDR contains many more up-to-date estimates: the latest year of data in ICDR is more recent than both GBD and UNSD for 105 countries, such as Algeria, Azerbaijan and Peru.^4^. Directly comparing ICDR estimates to existing ones reveals some surprising results. Algeria’s is estimated at 90% completeness, while UNSD and GBD estimate completeness at “Less than 90%” and 31% respectively and both estimates are outdated - from 2001 and 2006. Bolivia is estimated at 65%, while UNSD and GBD estimate completeness at “Less than 90%” and 47%, respectively. Again, both estimates are outdated - from 2000 and 2003. Bhutan’s vital statistics report for 2019, released on 2021, is the first one it issues, thus this is the first estimate of its’ completeness completeness level that I’m aware of, which I estimate as 71% complete. We were able to obtain registered death counts from Indonesia, and we estimate the completeness at 46% while neither GBD nor UNSD have any estimate.

For most of Africa, we were unable to obtain counts of registered deaths, and they thus have no estimate in ICDR. For the countries we were able to obtain information on death registrations, some African countries have the lowest estimated registration completeness: Nigeria and Uganda with less than 1% completeness; Countries such as as Liberia, Morocco, Ivory Coast and Rwanda at less than 40% complete. ICDR’s estimate for Kenya is much higher than that from UNSD and for Botswana it is very similar. We estimate virtually complete death registration in Egypt, Tunisia, South Africa and few more. A surprising and possibly false result is the essentially complete rate for Namibia. This is probably due to an unreasonably low estimate for the annual number of expected deaths in all three sources. In fact, Namibia’s own NSO in its’ vital statistics report estimate the expected number of deaths for 2017 at over 25 thousand (Namibia Statistics Agency, 2019), which would yield a much more reasonable completeness rate of 76%. We further discuss this in the Limitations section.

Europe’s vital registration is essentially complete across the board, with Andorra’s completeness estimate stands out as the lowest in Europe, and in contrast to UNSD (2021) which estimate it at “90% or more”. This might be due to the long time since that estimate was derived (2005) or due to the difficulty in establishing a reliable estimate for the expected number of deaths in such a small country (about 78,000 residents), as suggested by WPP and GHE not including estimates for countries with populations smaller than 900,000. Another country that stands out is Ukraine, which we estimate at about be 90%. This might be due to the annexation of Crimea by Russia, such that deaths that occur there are not registered by Ukraine, but the expected number of deaths in all of Ukraine still consider it part of Ukraine.

The countries of the Persian Gulf - Iran (100%), Iraq (80%), Kuwait (73%), Qatar (53%), Bahrain (71%), United Arab Emirates (42%), Oman (63%) and Saudi Arabia (59%) have relatively low estimated death completeness rates, with the exception of Iran (complete coverage). This is also in contrast to UNSD (2021) which estimate it as complete (Qatar, Saudi Arabia and the UAE) or close to 90% (Bahrain, Kuwait and Oman) with roughly similar high completeness estimates from GBD (2020). This might be due to the large numbers and share of immigrant, temporary workers in these countries, which have a very different demographic composition than the rest of the population, especially in terms of age - which the expected death estimates from WPP, GBD and GHE might not be taking into account properly. Many of these countries provide separate death counts for citizens and non-citizens. Unfortunately, neither of the sources for expected number of deaths contain separate estimates by citizenship status in these countries, so the analysis in ICDR is conducted on the total number of deaths. In the rest of Asia, several countries completeness estimate is lower than 50%, such as Afghanistan, Bangladesh, Laos and Pakistan while several countries have completeness estimates higher than 90%, such as the Philippines, Thailand, Malaysia, Kazakhstan and Sri Lanka.

In the Americas, ICDR contains estimates for all countries with the exception of Haiti. North America’s vital registration is complete while several countries in Latin America have relatively low completeness rates with Honduras, the Dominican Republic and Bolivia having the lowest estimated completeness in the analyzed time period at less than 65%.

## 4 Limitations

This study has several limitations. For many countries we were unable to locate information on the number of deaths recorded by their vital registration systems. We have chosen to treat such instances as missing rather than at 0% completeness - since the data might exist but it is not shared. Some countries have completeness of death registration estimates as arises from surveys or census. These were not included as information on vital registration is essentially missing. We have tried to make sure that the mortality counts reported here are based on vital registration and not estimates from other sources, but it is possible that we have misattributed some.

A significant limitation is the uncertainty embodied in the expected number of deaths, as derived from WPP, GBD and GHE. The expected number of deaths involves a complex estimation method that relies on many demographic variables as input (Rao et al., 2020). Some gauge of the uncertainty in the expected number of deaths is the disagreement between the three sources. The ratio or difference between the highest and the lowest expected number of deaths is sometime substantial. For example, WPP estimates almost 10 million deaths in India 2019, which is 750 thousand or 8% higher than the estimate from GHE. In Botswana, GBD estimates 23 thousand deaths, which is almost double (78% higher) than the estimate from WPP (13 thousand). A partial remedy for this limitation is to use more local knowledge on demographic processes. In many countries, the NSOs have their own estimates of total/expected number of deaths. Collecting such estimates systematically is beyond the scope of this work, but a few examples are in order: Namibia’s estimate is much higher than all three sources, India’s estimate is much lower than all three and Botswana is slightly lower than the mean of the three. Some of the countries’ 2019 estimate may be low due to incompleteness of the death registration process, resulting in a substantial drop in completeness in 2019, such as in the Maldives. However, as these estimates are from 2021 reports, they may indicate a deficiency in the vital registration system with regards to timeliness or disruption in 2020-2021 due to COVID-19 (see section 5).

This study has considered only national level counts of registered deaths to derive national-level estimates. However, national level registration completeness may obscure significant heterogeneity within countries. For example, the completeness of vital registration across the states of India ranges from 100% complete in Delhi, Gujarat, Karnataka, Kerala and more all the way down to 52% complete in Bihar and 21% complete in Manipur according to India’s own civil registration report (Office Of The Registrar General, 2021). In some countries, vital registration is functioning only in some regions, such that the NSOs report these figures as the total known registered counts. For example, in Djibouti, only the capital region of Djibouti-Ville, which contains about 66% of the total population, reports registered deaths; In Benin, the civil registration report explicitly states that the number of registered deaths is available only for “some communities”, resulting in a completeness estimate of about 2.6% in 2019. In incomplete vital registration systems, differences in death completeness are known to occur on other dimensions such as Urban/Rural, Sex, Age, Income and more (Basu and Adair, 2021; Adair et al., 2021).

Deaths that occur outside health facilities remain a challenge for many vital registration systems. For example, Burkina Faso and Liberia’s figures only relate to deaths that occur in hospitals and basic health facilities.

Some of the countries analyzed here were or are under severe political difficulties or civil war, examples include Libya, Syria, Yemen. In these countries the registered number of deaths reflect both the toll of civil conflict and the fact that some regions are not reporting registrations to the central government. For example, during the entire period 2015-2019 there was no reporting of the number of deaths from the Aleppo governorate in Syria.

Reports on vital registration in the analyzed period may be forthcoming. Sources are checked regularly and they will be added to the dataset on the public repository at https://github.com/akarlinsky/death_registration when published as ICDR is a live dataset.

## 5 Summary and Outlook

In this study we have presented the International Completeness of Death Registration (ICDR) dataset, that contains the latest completeness of death registration in 181countries from all over the globe. Completeness of death registration is a key feature and input into vital registration performance estimates, monitoring of SDGs and proper governance. There are other important properties of vital registration systems in general and death registration, in particular, that we did not consider here. These include but are not limited to, the timeliness of registration, i.e. the amount of time that passes between occurrence of death and it’s registration; The share of deaths that are registered with cause-of-death information (WHO, 2021), sex and age (Mikkelsen et al., 2015); The share of deaths registered with proper attribution of cause of death and a low share of “garbage cause of death codes”; etc. For example, while Egypt’s death registration is complete, it has a high share of garbage codes (Iburg et al., 2020). These dimensions of vital registration should be the focus of more research, especially in countries which have achieved complete or close to complete registration. In countries with incomplete registration, emphasis should be on strengthening basic vital registration as a necessary first step.

The period I have analyzed here is five years prior to 2020, the year COVID-19 began to spread. The effect of COVID-19 and government mandated measures to combat its’ spread on vital registration systems and the completeness rate is yet unclear. On the one hand, many vital registration systems “were either disrupted or discontinued” (Centre of Excellence for CRVS Systems, 2021; Pacific Community, 2021) which might lower completeness rates in 2020 onward. On the other hand, the high excess mortality observed in Ecuador and Peru, where weekly mortality has stayed well above historical levels even at times of relatively low numbers of reported COVID cases and deaths (Karlinsky and Kobak, 2021) might suggest an upward shift in completeness rates as well, with governments putting more resources into monitoring both general and COVID mortality.

An additional dimension of vital registration and dissemination that should be emphasized is the frequency of the published data. Excess mortality estimates, crucial to our understanding of COVID-19 (Beaney et al., 2020; Islam et al., 2021; Karlinsky and Kobak, 2021; Adair et al., 2020) as well as future disasters and pandemics, requires high-frequency mortality data, such as weekly or monthly. High frequency data is crucial for monitoring the true extent of pandemics, epidemics and disasters. As for estimates, while annual number of registered deaths counts are better than no registration data, their application to excess mortality estimates is limited since many of these events do not conform to annual timelines or last for a long duration. It is vital that countries publish high-frequency mortality data in the future, preferably by date of occurrence, just as they report periodical estimates of unemployment, prices and various other measures.

## Data Availability

All data is available at https://github.com/akarlinsky/death_registration

## Acknowledgments

We would like to thank Daniel Palacios and Gabrielle Halim for their help in obtaining the death registration data for Honduras and Indonesia. We greatly benefited from helpful comments and feedback from members of the WHO-UNSD Technical Advisory Group on COVID-19 Mortality Assessment and participants in the European Association of Population Studes: Health, Morbidity and Mortality Working Group Prague workshop. Special thanks to Dmitry Kobak, William Msemburi, Ilya Kashnitsky, Patrick Gerland and Bernardo Lanza Queiroz for extensive comments and discussion.

## Footnotes

1 The source of the UNSD figure is from Bangladesh’s Sample Vital Registration Survey, derived as the product of the survey CDR and population count, making it essentially an *expected* rather than the registered number of deaths.

2 Taken from the medium fertility variant time series.

3 While this corrects for the data issue, the state issue remains. Moldova exerts no de-facto control over Transnistria and Serbia over Kosovo, such that joining them together does not allow proper assessment of their independent vital registration systems.

4 ICDR contains more up-to-date estimates than UNSD for 127 countries and 134 for GBD

